# Normal neurophysiologic parameters of the tibial nerve among adult healthy Sudanese population

**DOI:** 10.1101/2023.04.26.23289151

**Authors:** Ahmed O Noury, Mohammed Salah Elmagzoub, Ahmed Hassan Ahmed, Hussam M A Hameed

**Affiliations:** Department of physiology, Orotta College of Medicine and Health Sciences, Asmara, Eritrea; College of Applied Medical Sciences, Imam Abdulrahman Bin Faisal University, Saudi Arabia; Faculty of Nursing, Jazan University, Saudi Arabia; Faculty of Medicine, National Ribat University, Sudan

## Abstract

**Background:** Nerve conductive studies (NCS) are the most informative portion of the electrodiagnostic evaluation for peripheral neuropathy; NCS can be extremely useful both in localizing lesions and determining the pathological processes. Recently, there is increased interest in quality of normative values for each test result and compare should be done between normal and patients regarding specific disease. So, The aim of our study is to help establish NCS normative data of tibial nerve that could be applicable in the Sudanese population and can be used in neurophysiology departments around the country and to compare with those data published in the literature, and to survey the effects of Age, Gender, Height, Weight and Temperature on Nerve Conduction Study Parameters.

**Methods:** NCSs were performed in 210 tibial nerves of 105 adult healthy Sudanese subjects using standardized techniques.

**Results:** It was found that the tibial nerve component values as follows; the distal latency is 4.063±1.0533 m/sec, Proximal latency 13.238 ±1.7253 m/sec, Conduction velocity 48.58±5.634 m/sec, F wave 51.509±6.1013 m/sec, Duration of CMAP at ankle 6.199± 1.0254 m/sec, Amplitude of CMAP at ankle 8.528±3.6658 μV, Area of CMAP at ankle 17.569±7.7389μV/ms, Duration of CMAP at popliteal 7.075±1.3173 m/sec, Amplitude of CMAP at popliteal 6.343±3.2871μV and Area at popliteal 14.914±7.8684 μV/ms.

**Conclusion:** The tibial motor nerve conduction parameters for the tested nerve compared favorably with the existing literature with some discrepancies that were justified

**Significance:** This is the first study to establish reference values for tibial NCS carried out Sudanese population.

## Introduction

Nerve conduction studies (NCS) are an important noninvasive diagnostic tools in the assessment of peripheral nervous system disorders and helps in diagnosis, prognosis, and monitoring of a disease process ^(1)^. NCS is diagnostically helpful in patients suspected of having almost any kind of peripheral nervous system disorder including disorders of nerve roots, peripheral nerves, muscle, and neuromuscular junction ^(2)^ Still there is limited published data and there is paucity of studies addressing NCS values of the common cranial, upper and lower limbs nerves in this region, including Sudan. Hence, this study aims to establish a normal Sudanese NCS reference values among adult healthy population. This will allow avoiding the inconsistencies that might be induced by many factors such as ethnic and environmental influences ^(3,4)^. The tibial nerve is one of the two terminal branches of the sciatic nerve, the largest nerve in the human body. The tibial nerve originates from the L4-S3 spinal nerve roots and provides motor and sensory innervation to most of the posterior leg and foot ^(5)^. It runs downward through the popliteal fossa, lying first on the lateral side of the popliteal artery, then posterior to it and finally medial to it. The popliteal vein lies between the nerve and the artery throughout its course. The nerve enters the posterior compartment of the leg by passing beneath the soleus muscle ^(6)^.

## Subject and methods

This is descriptive, cross-sectional non-interventional, clinic-based analytic, study conducted in El Magzoub Neuroscience Center and the Faculty of Medicine, National Ribat University, Khartoum, Sudan. NCSs were performed in 210 tibial nerves of 105 adult healthy Sudanese volunteers, selected after clinical evaluation to exclude systemic or neuromuscular disorders. Standardized techniques were used. The nature and procedures of the study were explained to volunteers. A verbal consent was obtained from each volunteer before the test and confidentiality was maintained. Some variables were obtained from the subjects by pre coded check list; height in centimeter; weight in kilogram; body mass index: calculated by the formula (Kg/m^2^) and body temperature in degree centigrade recorded immediately before performing the test. The study was performed at an ambient room temperature of 25°C with the subject lying comfortably in a supine position. A Viking Select Electromyography Machine was used and set at a low frequency cut filter of 2–5 Hz and the high frequency cut of 10 KHz. A standardized technique was used to obtain and record compound muscle action potentials (CMAPs) for motor studies. Both tibial motor nerves were stimulated orthodromically with bipolar surface stimulating electrode at two points along its course. Distal stimulation was behind and above the medial malleolus and proximally at the level of the knee in the lower border of the popliteal fossa near the popliteal artery. The active recording electrode is placed over the abductor hallucis muscle, the inactive electrode is placed on the muscle tendon at the level of the big toe. The ground is placed on the dorsum of the foot. CMAPs distal and proximal latencies, duration, area, amplitude, conduction velocity (CV), and F-wave were calculated.

### Statistical Analysis

Analysis was done using statistical package for social sciences (SPSS) 10.0 version. Values obtained were expressed in the form of mean and standard deviation (SD). P value was taken as significant if found to be <0.05. Comparison between right and left tibial nerves parameters was attained.

## Results

One hundred five volunteers with mean age=36.26 ± 9.978 were included in the study. The gender distribution and other anthropometric variables including BMI, height and body temperature were arranged into groups. The data obtained from nerve conduction study was suitably arranged into tables for discussion under different headings. Descriptive statistical analysis was carried out on this data and the results are presented as mean ± standard deviation and shown in table (1). The conduction finding of the whole tibial nerves and effects of gender on nerve conduction parameters is presented in Table (2). The mean and standard deviation for the right and left Tibial nerves CMAPs are summarized in Table (3,4). Table (5) shows a comparison between the results of the current study and those reported in other EMG laboratories.

**Table (1).** Descriptive Statistics of the study group

| Parameters | Minimum | Maximum | Mean | SD |
| --- | --- | --- | --- | --- |
| Age | 22 | 62 | 36.26 | 9.978 |
| **High** | 148 | 190 | 170.87 | 8.405 |
| **Body mass index** | 15.1 | 34 | 24.225 | 4.6281 |
| **Body temperature** | 34 | 37.8 | 36.162 | 0.7851 |

**Table (2).** Effects of sex on Tibial nerve conduction parameters.

| Site of stimulation | Parameter | | Distal Latency<br>(m/sec) | Duration<br>(m/sec) | Area<br>(μV/ms) | Amplitude<br>(μV) | Conduction<br>Velocity ( m/sec) | F.wave<br>(msec) |
| --- | --- | --- | --- | --- | --- | --- | --- | --- |
| Ankle | Male | Mean | 4.245 | 6.302 | 18.134 | 8.842 | 47.74 | 53.286 |
|  | N 130 | S.D | 1.0334 | 1.2277 | 6.8807 | 3.1871 | 5.666 | 4.3801 |
|  | Female | Mean | 3.768 | 5.960 | 16.740 | 8.645 | 49.73 | 50.905 |
|  | N 80 | S.D | 1.0304 | .9647 | 7.3749 | 3.6879 | 6.801 | 5.3121 |
|  | Whole sample | Mean | 4.063 | 6.171 | 17.603 | 8.767 | 48.50 | 52.379 |
|  | N 210 | S.D | 1.0533 | 1.1421 | 7.0706 | 3.3705 | 6.168 | 4.8722 |
| Popliteal | Parameter | | Proximal<br>latency (m/sec) | Duration<br>(m/sec) | Area<br>(μV/ms) | Amplitude<br>(μV) | Distance<br>(mm) | |
|  | Male | Mean | 13.631 | 7.355 | 15.055 | 6.555 | 448.92 |  |
|  | N 130 | S.D | 1.6620 | 1.9089 | 6.7022 | 2.8098 | 22.921 |  |
|  | Female | Mean | 12.768 | 6.858 | 12.655 | 5.752 | 434.75 |  |
|  | N 80 | S.D | 1.9410 | 1.3856 | 6.3925 | 2.8856 | 20.753 |  |
|  | Whole sample | Mean | 13.302 | 7.166 | 14.141 | 6.250 | 443.52 |  |
|  | N 210 | S.D | 1.8138 | 1.7382 | 6.6587 | 2.8521 | 23.079 |  |

**Table (3).** Motor conduction finding of the Right Tibial nerve

| Site of stimulation | Parameters | N | Minimum | Maximum | Mean | SD |
| --- | --- | --- | --- | --- | --- | --- |
| At Ankle | Duration(m/sec) | 105 | 3.1 | 9.5 | 6.171 | 1.1421 |
| | Amplitude( μV) | 105 | 2.6 | 18.0 | 8.767 | 3.3705 |
| | Area (μV/ms) | 105 | 4.6 | 33.8 | 17.603 | 7.0706 |
|  | Distal Latency (m/sec) | 105 | 1.9 | 6.7 | 4.063 | 1.0533 |
| At popliteal | Duration (m/sec) | 105 | 3.5 | 16.4 | 7.166 | 1.7382 |
| | Amplitude(μV) | 105 | 1.2 | 15.0 | 6.250 | 2.8521 |
| | Area(μV/ms) | 105 | 1.4 | 33.1 | 14.141 | 6.6587 |
|  | Proximal latency (m/sec) | 105 | 6.9 | 17.2 | 13.302 | 1.8138 |
|  | Distance( mm) | 105 | 390 | 500 | 443.52 | 23.079 |
|  | Conduction velocity( m/sec) | 105 | 37 | 67 | 48.50 | 6.168 |
|  | F wave (m/sec) | 105 | 33.5 | 64.3 | 52.379 | 4.8722 |

**Table (4).**
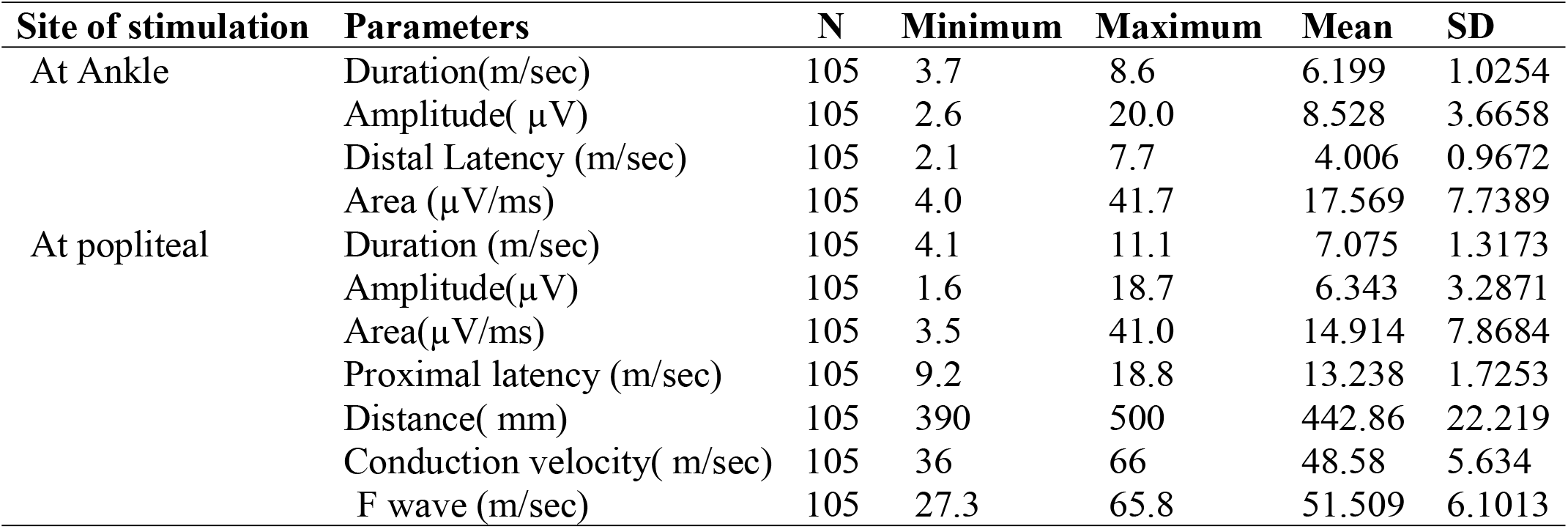
Motor conduction finding of the Left Tibial nerve.

| Site of stimulation | Parameters | N | Minimum | Maximum | Mean | SD |
| --- | --- | --- | --- | --- | --- | --- |
| At Ankle | Duration(m/sec) | 105 | 3.7 | 8.6 | 6.199 | 1.0254 |
| | Amplitude( μV) | 105 | 2.6 | 20.0 | 8.528 | 3.6658 |
|  | Distal Latency (m/sec) | 105 | 2.1 | 7.7 | 4.006 | 0.9672 |
| | Area (μV/ms) | 105 | 4.0 | 41.7 | 17.569 | 7.7389 |
| At popliteal | Duration (m/sec) | 105 | 4.1 | 11.1 | 7.075 | 1.3173 |
| | Amplitude(μV) | 105 | 1.6 | 18.7 | 6.343 | 3.2871 |
| | Area(μV/ms) | 105 | 3.5 | 41.0 | 14.914 | 7.8684 |
|  | Proximal latency (m/sec) | 105 | 9.2 | 18.8 | 13.238 | 1.7253 |
|  | Distance( mm) | 105 | 390 | 500 | 442.86 | 22.219 |
|  | Conduction velocity( m/sec) | 105 | 36 | 66 | 48.58 | 5.634 |
|  | F wave (m/sec) | 105 | 27.3 | 65.8 | 51.509 | 6.1013 |

**Table (5).** A comparison CMAP values of the Tibial nerve between the current study and some reported in other EMG laboratories.

| **Studies** | **DML (msec)** | **CMAPA (μV)** | **CV (m/sec)** | **F.wave (msec)** |
| --- | --- | --- | --- | --- |
| Present study<br>N = 105 | 4.063±1.0533 | 8.767± 3.3705 | 48.50 ± 6.168 | 52.379±<br>4.8722 |
| Vijay(Haryana “northern India”) <sup>15</sup><br>N=120 | 4.1 ± 0.86 | 6.17 ± 0.43 | 46.95 ± 3.2 | ----- |
| Shahabuddin (Aurangabad“western India”) (7)<br>N=90 | 4.77 ± 0.36 | 6.22 ± 0.48 | 45.52 ± 3.04 | ----- |
| Ginanneschi (Italy) (8)<br>N= 185 | 4.52 ± 0.7 | 6 ± 2.5 | 44.3 ± 7.6 | ----- |
| Mukesh (Nebal) (9)<br>N=40 | 10.84±0.64 | 15.22±1.14 | ----- | 42.54±1.18 |
| Zaitoon (Pakistan) (10)<br>N=100 | 3.9±0.6 | 12.5±4.5 | 48.6±4.1 | 45.4±4.1 |
| Amal (Egypt) (11)<br>N=100 | 3.4±0.6 | 8.6±2.9 | 49.4±3.3 | 44.9±3.8 |
| Farqad (Irag) (12)<br>N = 11,437 | 4.09±0.63 | 8.97±4.45 | 49.59±5.26 | 49.74±4.87 |
| Chi-Ren(Taiwan) (13)<br>N = 101 | 4.0 ±0.7 | 16.5 ± 4.7 | 47.5 ± 2.8 | 46.0 ± 2.9 |
| Rasheedi(Saudi) (14)<br>N=76 | 3.5±0.61 | 12.1±3.30 | 48.7±4.6 | ----- |

The values are presented as mean ±SD, SL; CV (conduction velocity); DML (Distal motor Latency); CMAP (compound muscle action potential amplitude)

Two hundred ten tibial nerves of 105 healthy Sudanese subjects were included in the study. Sixty-two (62%) were males and thirty-eight (38%) were females. Their ages ranged between 22 and 62 years with an overall average of 36.22 ± 9.978 years. Most of them (67%) were within the age bracket of 22 and 39 years. They have an average weight of 71.04 kg, height of 170.97cm and average BMI 24.225 (Table1). The recorded CMAP mean ± standard deviation (with range between brackets) values in the whole study group (210 nerves) were found to be as follows: at ankle stimulation; distal latency, 4.063±1.0533 ms (3-5.1); amplitude, 6.250±2.8521 mV (3.4-9.1); duration, 7.166±1.7382 ms (5.4-8.9); and area 17.603 ± 7.0706 mV/ms (10.5-25.3). Values at popliteal stimulation; proximal latency, 13.302 ± 1.8138ms (11.48-15.1); amplitude, 8.767 ± 3.3705mV (5.4-12.1); duration,6.171±1.1421ms (5.0-7.3); and area 14.141± 6.6587 mV/ms (7.49-207). Conduction velocity, 48.58 ± 6.168 m/s (42.4-54.7); and F wave 52.379 ± 4.8722ms (47.5-57.2) (Table 2). The right and left tibial nerves values in the whole study group were presented tables 3 and 4, while the effect of gender and side of tibial nerve parameters were shown in table 5 and 6. Table (7) shows a comparison between the tibial nerve parameters of the current study and those reported in literature.

## DISCUSSION

This is the fourth article of a series of NCSs of the common upper and lower limb peripheral nerves among healthy adult Sudanese people (16–18); aiming to establish our own normative reference values of tibial nerve for our EMG laboratories in Sudan. The study also aimed to investigate the effect of age, gender, high, BMI and temperature on NCS variables in healthy adults.

A comparison was made between this study and other studies published in literature; the comparison of our finding and other studies of normative neurophysiological data in different countries were presented in the Table (5). In table (5), it can be observed that the CV of the tibial nerve was favorable with the results of Zaitoon (10) Rasheedi (14)the CV were higher than that reported by Shahabuddin (7) Ginanneschi (8)and Shehab (15). On the contrary, the CV was less than that reported by Amal (11) and Farqad (12). Regarding CMAPA values were similar to those encountered by Amal (11)and Farqad (12)On the other hand CMAPA the were higher than that reported by Vijay^(15)^ Shahabuddin (7)and Ginanneschi (8)On the contrary, the CMAPA was less than that reported by Chi-Ren (13)Rasheedi (14)Zaitoon (10)Mukesh (9). The values of F-wave parameter recorded from the tibial nerve were highest than that reported by others

Age has been widely accepted to have an influence on nerve conduction parameters as noted in literature(19,20) Aging process is associated with nerve degeneration; older subjects have longer distal latency, smaller amplitude and slower nerve conduction velocity than younger subjects as noted by Stetson et al.(20) and Mayer et al (21). In this study we found that the distal latency of Tibial nerve increases with age A similar association between increasing age and increasing distal latency has been noted by Saufi et al (22)Stetson et al (20)and Mayer et al (21).In our study, the CMAP amplitudes (ankle and popliteal) decreases with age. In agreement with our study, Huang et al reported that age factor had been negatively correlated to amplitude (13). In this study the conduction velocity of Tibial nerve significantly decreases with age. A similar finding was observed by Kumar et al (23) and Saufi et al (22) in their study found age was found to have a negative correlation with conduction velocity. Suchitra et al (3) found that F-wave was positively correlated to age, and the same is proved in our study. In summery older subjects have longer distal latency, smaller amplitude, slower nerve conduction velocity and longer F wave than younger subjects. Probably, the reason behind these findings the aging process is associated with nerve degeneration

This paper reviews a number of studies on the correlation between gender and nerve conduction study parameters, gender have a significant effect on amplitude, duration and latency of motor nerve conduction studies as reported by Huang et al (13) and Sajadi et al (24). In the present study, we found that all Tibial nerve conduction study parameters are less in female in comparison to male except the conduction velocity is significantly higher in female which are similar the findings Stetson et al (20) Dilip et al (25). Also, in our study, F-wave latencies of Tibial nerve were longer in males as compared to females and the probable reason is greater height and limb length in male volunteers. Our results disagree with study of Huang et al (13)and Wasim et al (26) who revealed that F-wave latencies in both Tibial nerves were greater in females

A significant negative correlation between height and nerve conduction velocity and positive significant negative correlation between the height and F wave latency was also observed. Our findings in agreement with earlier reports that report an inverse relationship between height and nerve conduction velocity and direct relation between the F wave latency (15,18,20).

The increased BMI is found to be associated with lower amplitude both ankle and popliteal CMAP. Similar were the findings reported by Pawar et al (27). This observation might be due to amplitude lessening by thicker subcutaneous tissue in the person with higher BMI (28). In our study, Increasing BMI is associated with prolongation of distal motor latency these findings is consistent with the finding of Pawar et al (29)The conduction velocity slow with increasing BMI Our observation are in agreement with Pawar et al (27)and Awang MS et al (22) who observed slowing of conduction velocity with increasing BMI. F-wave minimum latency showed decrease with increasing BMI Similar were the findings reported by Pawar et al (27). Also, contrary to our study, Baqai HZ et al (29)reported no effect of BMI on nerve conduction studies.

This is a preliminary result, further study recruiting large number of participant is recommended.

## Conclusion

NCS values were well established in the Western countries several decades back and now unfortunately, are used as reference values in many EMG laboratories across the world, including our region; regardless of the environmental and ethnic discrepancies. Since NCSs were found to be one of the most sensitive tests in the diagnosis of peripheral nerves pathologies and NMJDs, it became extremely important to set local normal baseline electrophysiologic parameters for these nerves. In this study series normative values of tibial nerve were established in the Sudanese population. Overall, motor nerve conduction parameters for the tibial nerve compared favorably with the existing literature and appeared to be influenced by gender, height, and BMI.

## Data Availability

All data produced in the present study are available upon reasonable request to the authors

## Recommendations

Apart from what has been achieved from this study, it is recommended to extend the study to include other EMG labs in Sudan that are reliable and to study all the nerves of the upper and lower limbs.

## Acknowledgments

We acknowledge the technical support of Ustz. Kholoud Abudaif for her help in the data analysis and interpretation. Dr. Kamal Mohamed for providing statistical assistance

## Conflicts of interest

There are no conflicts of interest.

## Notes

### Competing Interest Statement

The authors have declared no competing interest.

### Funding Statement

This study did not receive any funding

### Author Declarations

The National Ribat University ethics committee granted ethical approval for this study.

